# Interruption Managements Skills: Identifying and Addressing the Learning Needs of Medical Students: A Protocol for an Interview-based Study

**DOI:** 10.1101/2025.03.11.25323751

**Authors:** Assim Javaid, Michal Tombs, Steven Riley, Philip Morgan

## Abstract

**Background and objectives:** Interruptions during clinical activities are strongly associated with patient safety incidents in healthcare settings. Attempts have been made to teach medical students and doctors how to best manage interruptions, however evidence suggests that most doctors develop these skills through experience and that students feel ill-prepared when confronted with interruptions in the clinical setting. This study protocol presents a study that aims to inform, the development of an educational tool for undergraduate medical students on management on interruption management in the clinical setting. Informed by the ADDIE model of instructional design, a learning needs analysis will be conducted to determine what skills are needed to best manage workplace interruptions and how these skills would best be taught at an undergraduate level.

**Methods and Analysis:** This study will collect data through semi-structured interviews with medical students, medical educators and junior doctors. The interview protocol has been developed using the Critical Incidence Technique, in which real-life experiences will be used as a basis for reflection to determine what underlying lessons can be learnt from them. The interviews will be transcribed verbatim and undergo thematic analysis to determine what instructional goals need to be set for an educational tool and how such a tool should be designed.

**Ethics and Dissemination:** This study has received full ethical approval from Cardiff University (SREC reference 23/67). Findings from the study will be disseminated by presentation at national and international conferences and publication through academic papers.

## Introduction

Literature on the link between interruptions in clinical settings with patient safety incidents (Monteiro et al, 2015; McMullan et al, 2021; Balint et al, 2014; Kellogg et al. 2021) highlights the way by which interruptions interfere with the decision-making process (Finn et al, 2008). Whilst interruptions do overlap with distractions, they are in fact theoretically different. Distractions, referred to by Odberg et al (2018) as “passive interruptions”, are events that have the potential to reduce concentration but do not disrupt work (e.g., music playing in the background). This is differentiated from interruptions as these events cause the primary task to be disrupted. Odberg et al (2018) referred to these events as “active interruptions” and a key element of them is the deliberateness of the interrupting task, with a conscious effort being made by a third part to draw an individual’s attention from their primary task (e.g., a direct question from a member of staff).

Research suggests that it may prove difficult to eliminate all interruptions from a healthcare environment because of its inherent dynamism, however, there may be a role in helping healthcare staff develop skills in managing the deliberate interruptions that they will face (Hayes et al. 2015). Johnson and colleagues examined the efficacy of using e-learning modules to teach nurses preferred behavioural strategies for managing interruptions. Nurses were told to preferentially continue with their original task by blocking any attempts at interruption (Johnson et al. 2018a; Johnson et al. 2019). They were also advised not to attempt to multitask. They found, however, that, when observed, these nurses were no more likely to use the preferred strategies than a control group who were not taught the strategies. When discussing this in focus groups, the nurses felt the teaching would lead to some behavioural change but that interruptions were inevitable. In contrast, evaluation studies conducted in simulation settings did find educational interventions to be effective. Two examples come from studies where participants were taught the Stay S.A.F.E. method for interruption management. This method involves five stages to manage interruption and is based on the Memory of Goals Theory (Altmann and Trafton, 2002), which proposes that using priming and goal-directed cues allows individuals to better manage interruptions. Both studies used a PowerPoint presentation to teach this tool to either nurses (Henneman et al. 2018) or nursing students (Vital and Nathanson 2023), and both found reduced distraction time during simulated clinical scenarios.

To date, most published studies on the topic of teaching interruption management skills were conducted in simulated settings. In these studies, learning largely focused on raising awareness of the reality of interruptions and distractions in the workplace in order to encourage the participant to identify their own learning points. The learning was either self-directed through reflective practice (Hayes et al. 2017; Hayes et al. 2018; Hayes et al. 2019; Nowell, 2023) or guided during debrief (Thomas et al, 2014; Thomas et al, 2015; Thomas, 2015). Thomas et al (2015) found that medical students’ who were given personalised feedback about distraction and interruption after a simulation had a significantly greater improvement in error rates than a control group. These studies have found that participants reported feeling more confident about their skills or enjoying the simulations.

### Purpose

Given the risks to patient safety, equipping medical students with basic interruption management skills could have significant benefits. However, the existing literature does not provide much clarity regarding what should be taught and how. Very few of the studies on the topic described the way in which their educational interventions were developed, lacking the detail regarding pedagogy and educational theory. In particular, it was not clear whether a learning needs assessment was conducted before developing the intervention and the extent to which educational literature and pedagogy informed the development of these. This study attempts to address this gap by developing an educational intervention that is is informed by the ADDIE (Analysis, Design, Development, Implementation, Evaluation) instructional design model (Aldoobie, 2015)starting with a comprehensive learning needs analysis The first step of the ADDIE model is Analysis. In this phase we need to address what the learners already know and what their learning needs are (Aldoobie,2015). A useful means to complete this aspect is through a “learning needs assessment” (Grant 2002), which provides frameworks through which educational needs can be determined. One approach to a learning needs assessment is the use of “learner-identified needs” (Pilcher, 2016), where interviews can be used to help individuals reflect on and identify their own learning needs. As this project is aimed at helping medical students learn how to manage interruptions, it is key to interview them in order to ensure their own self-identified needs are met. Another aspect of the Analysis stage of the ADDIE model is the identification of instructional goals (Aldoobie, 2015), which is to say identifying what it is that the learners need to learn. In the context of learning a non-technical skill, a useful approach to this is through “Reflection on Action” (Grant, 2002), in which those who have relevant experiences reflect back on those experiences to identify what went well and what could have gone better. Interviewing junior doctors and asking them to reflect on experiences of interruptions will aid in identifying these instructional goals. Similarly, interviewing medical educators with experience in teaching non-technical skills may offer insights into instructional goals that junior doctors or medical students may not have considered.

### Overall aim and Research Questions

This study will conduct a comprehensive needs analysis from the perspective of three main stakeholders including junior doctors, medical students and medical educators. The specific research questions to be addressed are:

1. What are participants’ experiences of learning interruption management skills in the clinical setting?
2. What training did participants receive for managing interruptions?
3. What educational approaches would be best suited to aid the learning of these skills for medical students?

### Anticipated outcome

This learning needs analysis will inform the design and development of an educational intervention on teaching interruption management skills for Undergraduate Medical Students in the UK.

## Methods and Analysis

### Design

The study will use the Critical Incident Technique (CIT) (Flanagan, 1954) to determine themes around interruption management skills CIT involves asking participants to recall specific events relevant to the research question (e.g., a time they learned about managing an interruption in the clinical setting) and uses this event as a starting point to explore what they gained from the experience and the impact it had on their behaviour (Tombs, 2019). The focus is on lessons learned from the experience, rather than the details of the experience in itself. This is a particularly useful technique for exploring these skills as participants are very likely to have direct or observed experience of interruptions and may have even been taught, or will themselves have taught, these skills. The CIT also ensures both positive and negative experiences are explored by switching between the two types of experiences following each incident explored.

## Participants and recruitment method

Participants will be recruited via social media. Social media platforms will include X (formally known as Twitter), LinkedIn and WhatsApp. Invitations being disseminated openly on public platforms (X and Linkedin) and via personal networks (through WhatsApp).

Further recruitment will occur with the snowballing technique (Browne, 2005), where those who have seen the initial invitation pass details of the study on to those they know with similar interests, allowing for a wider area of distribution of the invite. The invitation will include a link to an online Participant Information Leaflet, which will give potential participants more details about the study, allowing for informed consent. They will then be prompted to complete an online consent form if they are happy to be involved with the study.

Inclusion criteria:

- any medical student who is currently studying medicine in the UK or doing additional studies as a temporary, planned break from their medical degree;
- junior doctors including all foundation years doctors, those in training posts and those working in a non-training post, where the level of clinical responsibility is not more than those in a training post;
- medical educators who are involved in the teaching of either medical students or junior doctors in any clinically related capacity.

Exclusion criteria:

- medical students, junior doctors or medical educators who are not from the UK;
- junior doctors who are not working clinically as a doctors.

In line with qualitative research methodology, participants will continue to be recruited until data saturation is achieved and no new themes are forthcoming during the interviews (Glaser and Strauss, 1973). It is estimated that a total of 30-60 participants will be adequate for this study; that is to say, 10-20 participants from each group: medical students, junior doctors, and medical educators – noting the points on saturation above.

### Materials and Data Collection

CIT will be used to generate a semi-structured interview template. The interview structure will be altered to make it more appropriate for the group being interviewed. For example, a line of questioning for junior doctors will be:

Can you think of a few examples of when you have found interruptions to be useful. I’d like to explore each of these examples in turn.

1. Thinking of one of these examples, can you describe the circumstances of the interruption?
2. What made this a positive experience?
3. What did you learn from this experience? What impact did this, or will this, experience have on the way you do things?

To avoid order effects, the requests for a positive and negative experiences will be counterbalanced, with some participants being asked to relay positive incidents first and others being asked to start with negative incidents.

The first two interviews in each stakeholder group will be used as pilots to refine the questions to make them more appropriate for future interviews if this is needed. These initial interviews will be included in the overall dataset for analysis. Interviews will be conducted and recorded online using Microsoft Teams

### Data Analysis

This study will follow the SRQR guidelines for reporting qualitative research (O’Brien et al, 2014). Interviews will be transcribed verbatim, then analysed using the principles of thematic analysis (Braun and Clarke, 2022). Inter-rater reliability will also be calculated (Tombs, 2019).

### Patient and Public Involvement

This study is part of a PhD in Medical Education, for which the initial proposal received feedback from a Public and Patient Involvement (PPI) group, funded by Health Care Research Wales. This helped shape the direction and priorities of the PhD as a whole. Feedback on the project proposal was sought from seven lay members of the public. All felt that this would be a useful project. Specific feedback that was factored into the project design was to try to target underrepresented groups of the population if the initial call for participants did not garner enough diversity, to use social media platforms to aid in getting a more diverse group of participants, and to consider sharing findings with non-medical healthcare professionals also.

### Ethics and Dissemination

This study has received full ethical approval from Cardiff University (SREC reference 23/67). Findings from the study will be disseminated by presentation at national and international conferences and publication through academic papers.

## Supporting information

Ethics Approval

## Data Availability

There was no data used for this manuscript as it is a study protocol. The Ethics Committee gave approval for the proposed study to go ahead.

## Authorship

AJ and MT were responsible for conceptualisation and for writing the protocol and initial few drafts. All authors were responsible for writing subsequent drafts and revisions. AJ was responsible for study design and data collection with supervision from MT, SR and PM.

