## Supplementary material for "Interruption Managements Skills: Identifying and Addressing the Learning Needs of Medical Students: A Protocol for an Interview-based Study": Ethics Approval

School of Medicine
Yr Ysgol Meddygaeth

Cardiff University
Main Building
Heath Park
Cardiff CF14 4XN
Wales, UK
Prifysgol Caerdydd
Prif Adeilad
Parc y Mynydd Bychan
Caerdydd CF14 4XN
Cymru, Y Deyrnas Unedig

14th November 2023

JAVAID, Assim

Centre for Medical Education

Dear Assim,

**Research project title:**

Interruption Managements Skills in Medical Education: A Qualitative Study to Identify Learner Needs

**SREC reference:**

23/67

The School of Medicine Research Ethics Committee ('Committee') reviewed the above application at the meeting held on 13th September 2023.

**Ethical Opinion**

The Committee gave:

A a favourable ethical opinion of the above application on the basis described in the application form, protocol and supporting documentation.

**Additional approvals**
This letter provides an ethical opinion only. You must not start your research project until all appropriate approvals are in place.

**Amendments**
Any substantial amendments to documents previously reviewed by the Committee must be submitted to the Committee via email to Dr Michael Laing and Mr Konstantinos Grigoratos for consideration and cannot be implemented until the Committee has confirmed it is satisfied with the proposed amendments.

You are permitted to implement non-substantial amendments to the documents previously reviewed by the Committee but you must provide a copy of any updated documents to the Committee via email to for its records.

**Monitoring requirements**
The Committee must be informed of any unexpected ethical issues or unexpected adverse events that arise during the research project. In addition to this, the Committee request an end of project report sent to the Committee via email to. This must be sent along with confirmation that your research project has ended and sent within the three months of the research project completion.

**Documents reviewed by Committee**
The documents reviewed by the Committee were:

| Document | Version | Date |
| --- | --- | --- |

**Complaints/Appeals**

If you are dissatisfied with the decision made by the Committee, please contact the Chair of the Committee via the Committee Secretary in the first instance to discuss your complaint. If this discussion does not resolve the issue, you are entitled to refer the matter to the Head of School for further consideration. The Head of School may refer the matter to the Open Research Integrity and Ethics Committee (ORIEC), where this is appropriate. Please be advised that ORIEC will not normally interfere with a decision of the Committee and is concerned only with the general principles of natural justice, reasonableness and fairness of the decision.

Please use the Committee reference number on all future correspondence.

**The Committee reminds you that it is your responsibility to conduct your research project to the highest ethical standards and to keep all ethical issues arising from your research project under regular review.**

**You are expected to comply with Cardiff University's policies, procedures and guidance at all times, including, but not limited to, its Policy on the Ethical Conduct of Research involving Human Participants, Human Material or Human Data and our Research Integrity and Governance Code of Practice.**

Yours sincerely,

Dr Ned Powell
Chair, School of Medicine Research Ethics Committee

Registered Charity No. 1136855
Elusen Gofrestredig Rhif. 1136855
